# Pathway-based genetic susceptibility and cleaning agent exposures in adult asthma: A semi explorative GxE analysis in the Personalized Environment and Gene Study (PEGS)

**DOI:** 10.1101/2025.06.29.25330505

**Authors:** Yujing Chen, Ming Kei Chung

## Abstract

**Background:** Pathway-based approaches may effectively dissect the polygenic architecture of complex diseases. We conducted a semi-explorative interaction study between genome-wide and pathway-specific polygenic risk scores (PRS) and occupational asthmagens (i.e., cleaning agents) on adult asthma.

**Methods:** This study included 2615 adults from the Personalized Environment and Genes Study (PEGS) in North Carolina, with comprehensive questionnaire-based health and exposure data and whole genome sequencing data. Occupational exposure to any cleaning agent was assessed as ever exposure to ammonia, chlorine bleach, and carbon tetrachloride over 15 minutes per week in any job. Current adult asthma was defined as doctor-diagnosed asthma with an attack in the past year. We estimated genome-wide PRS and pathway-specific PRS of oxidative stress and type 2 immune responses. We used 1) logistic regression to analyze interactions between PRS and exposure, and 2) linear principal-component regression to examine gene-based interactions.

**Results:** Current asthma prevalence was 9.56% (n=250). Occupational exposure to cleaning agents was associated with an increased asthma risk [adjusted odds ratio (aOR): 1.56-2.24]. Genome-wide PRS multiplicatively interacted with any cleaning agent or bleach (*P* _interaction_ =0.029 or 0.039), while pathway-based PRS conferred asthma risk independently. Additive interactions between cleaning agents and high genetic risks (PRS>median splits) were observed, showing the highest excess asthma risk in individuals with both exposure and pathway-based risks. Notably, eight genes were identified to explain oxidative stress PRS-related interaction with cleaning agents on asthma (*P* _interaction_ < 0.05/29 genes).

**Conclusion:** Applying novel and statically powerful pathway-based PRS, we found cleaning agents synergistically interacted with genetic risks of asthma, advancing mechanistic insights for precision prevention and intervention in asthma.

## Introduction

Asthma is a heterogeneous chronic inflammatory disease, affecting approximately 7.7% of the U.S. population, including over 20 million adults. ^1^ Compared to childhood-onset asthma, adult-onset asthma is often less atopic and more symptomatic, with occupational exposures serving as important triggers.^2^ Cleaning products are common asthmagens, which contain a mixture of chemical irritants and sensitizers and can cause excessive exposure to other toxicants, such as volatile organic compounds (VOCs).^3^ Occupational cleaning product exposures have increased over recent years across all industries ^4^ and were linked to exacerbated respiratory symptoms and new-onset asthma. ^5–7^ However, underlying mechanisms in relation with individual genetic profiles for cleaning agent-induced asthma risks remain poorly understood.

Genetic susceptibility also constitutes a substantial component of asthma. Twin studies estimate the heritability of adult asthma to range from 57% to 73%. ^8,9^ These genetic factors likely contribute to asthma through pathogenetic mechanisms—such as oxidative stress, airway inflammation, and type 2 cytokine-mediated immune response—that may involve interactions with exogenous oxidants and allergens. ^10–12^ Studies using the candidate gene approach have suggested that antioxidant and immune-related genes could modify the effects of air pollution and tobacco smoke on asthma. ^13–15^ However, this approach may not capture the polygenic architecture of complex diseases like asthma. ^16^ Recently, polygenic approaches, such as polygenic risk scores (PRS) have been developed to predict disease risk and explore gene-environment (G ⊆ E) interaction ^17^. The classical genome-wide PRS aggregates risk alleles across the genome into a single liability value but may be computationally intensive and risk information loss by obscuring individual genetic profiles and heterogeneous mechanisms of complex diseases. Hence, PRS estimated from specific functional pathways may offer a superior balance between statistical power and systematic dissection of functional structures of the genome. ^18^

Cleaning agents are thought to have oxidizing or sensitizing potentials ^19^, while comprehensive examinations of their interactions with aggregated genetic risk are lacking. Previous genome-wide interaction studies have relied on a single locus-based G ⊆ E approach for occupational exposure and respiratory outcomes. ^20,21,22^ Using pathway-based selection method, a European cohort study examined interaction between each oxidative stress-related gene with occupational exposure to low molecular weight (LMW) agents and irritants for adult-onset asthma. ^20^ Pathway-based PRS may serve as a more effective tool for studying G ⊆ E interaction and provide mechanistic insights specific to occupational cleaning agents.

Using an exploratory data-driven approach, our previous work identified cleaning agents as top asthmagens from 93 occupational chemical agents. ^23^ This study aimed to deepen our understanding of G ⊆ E interaction for adult asthma by specially (1) assessing asthma genetic risk with genome-wide and pathway-based PRS; and (2) analyzing multiplicative or additive interactions between occupational cleaning agents and PRS. We hypothesized that cleaning agents interacted with genetic susceptibility to adult asthma, particularly through oxidative stress and type 2 immune response pathways, due to oxidizing or sensitizing potentials of exposure.

## Methods

### Study population

The Personalized Environment and Genes Study (PEGS) is a multi-ancestry cohort established in 2002 at the National Institute of Environmental Health Sciences. Participants (≥18 years) were recruited through convenience sampling from outpatient clinics, hospitals, and the general population at over 200 unique sites in North Carolina. PEGS collected participants’ blood samples and administered three cross-sectional surveys to investigate extensive health conditions and environmental exposures from 2013 to 2022. As shown in **Figure 1 (A)** and **Figure S1**, 3230 adults had complete survey data, and 4737 underwent whole-genome sequencing (WGS) (version Freeze 3.1). We included 2660 participants with available environmental exposure, asthma, and eligible WGS data. After excluding those with missing values in age and race, the final analysis included 2615 adults. Local institutional review boards approved the study, and all participants provided written informed consent at the study enrollment.

**Figure 1.**
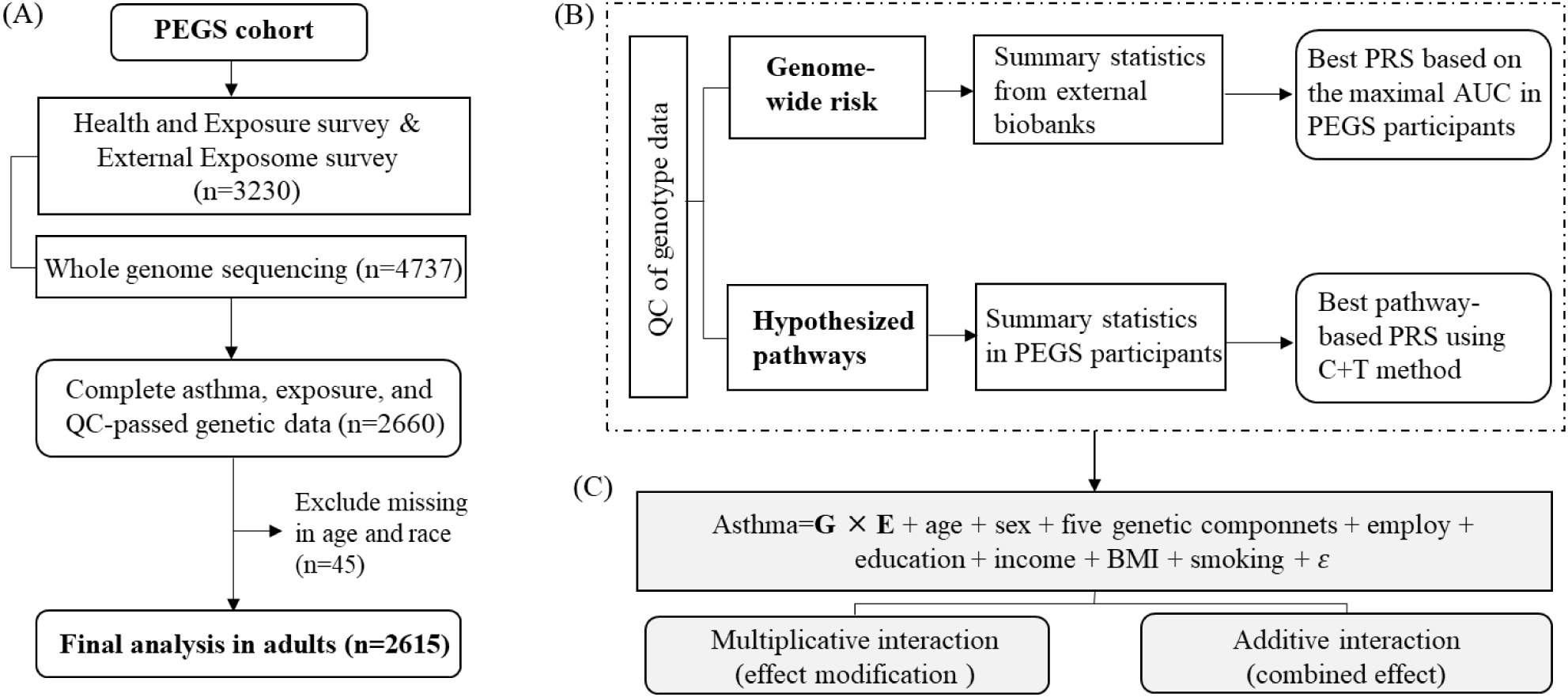
Study design and workflow. (A) The participants inclusion flowchart; (B) The development of genome-wide and pathway-based PRS for oxidative stress and type 2 immune response; (C) G and E interaction analysis. Abbreviation: PEGS, Personalized Environment and Genes Study; QC, quality control; AUC, area under the curve; PRS, polygenic risk score; C + T, clumping + thresholding; G: Genetic factor, referring to genome-wide and pathway-based PRS in this study; E: environment, referring to cleaning agent exposure in this study. Summary statistics of seven previously published genome-wide PRS were derived from UK Biobank (https://biobankengine.stanford.edu/prs) and Global Biobank Meta-analysis Initiative (https://www.globalbiobankmeta.org/resources). Pathway-based PRS were developed based on two curated gene sets (also see supplementary Figure S3).

### Genotyping and quality control of WGS data

Genotyping methods for PEGS samples have been detailed by Akhtari et al ^24^. Briefly, WGS was performed by the Broad Institute using the NovaSeq 6000 platform with a target genome-wide read depth of 30x. Samples aligned to the GRCh38 reference genome. Genome Analysis Toolkit (GATK) and DeepVariant were used to obtain variant calls and joint genotypes, resulting in over 43 million high-quality variants.

Quality control of the genome-wide SNPs was conducted using PLINK 1.9 (Harvard University). The analysis included only biallelic, autosomal single nucleotide polymorphisms (SNPs) that had passed PEGS initial quality control.^26^ We then filtered out SNPs with a call rate less than 95%, minor allele frequency less than 5%, and those failing the Hardy-Weinberg test at a 1.0⊆10^-4^ threshold. Samples were further excluded if they had a missing call rate > 5%, pi-hat >0.2 (indicating second-degree relatives), or deviating ±3 SD from the mean heterozygosity rate. The first five genetic principal components (PCs) of this multi-ancestry population were derived after pruning SNPs in high linkage disequilibrium. The pruning process resulted in 298,851 SNPs, using pairwise independence function with a 5-kilobase window shifted by 50 base pairs and an *r*^2^ < 0.2 between any pair.

### Occupational cleaning agent exposure

In the External Exposome Survey, exposure to cleaning agents was assessed, defined as contact through inhalation, skin contact, ingestion, or proximity to unsealed cleaning liquids for at least 15 minutes per week in the workplace. Participants were asked whether they had been exposed to ammonia / chlorine bleach / carbon tetrachloride /or other cleaning agents in any job they have held. For those exposed to any cleaning agents, information on exposure frequency (daily / weekly / yearly) and duration (years) at work was also collected.

### Asthma outcome assessments

In the Health and Exposure Survey, self-reported asthma diagnosis was determined by an affirmative answer to the question, “Has a doctor or other healthcare provider ever told you that you have asthma?” Current adult asthma cases were defined as those who 1) reported asthma diagnosis, and 2) still had asthma in adulthood or experienced either an asthma attack, emergency room visits, or received medication prescriptions in the past 12 months. Adult asthma was further classified as adult-onset and childhood-onset (adult exacerbated) by the diagnosis age (≥ or <18 years). Asthma related questions were adapted from validated questionnaires used in the Nurses’ Health Study II, NHANES, and the PhenX toolkit. ^27,28^

### Covariates

Demographic characteristics were collected through self-administered questionnaires in the Health and Exposure Survey. These included age, sex, race (White, Black, and others including Asian, American Indian or Alaskan Native, or multiple), highest education levels (high school or below, college, technical or vocational school graduate, and bachelor, and graduate or professional), annual household income (less than $20,000, $20,000 to 49,999, $50,000 to 79,999, $80,000 or above), employment status (currently employed or not), and smoking (smoked>100 cigarettes in a lifetime). Body mass index (BMI) was calculated based on self-reported height and weight and categorized into three groups (<25, 25-<30, and ≥30 kg/m^2^).

### Statistical analysis

#### General analyses

Participant characteristics were summarized as means and standard deviations for continuous variables or as frequencies and percentages for categorical variables. Differences between asthma and non-asthma groups were compared using *t*-tests or χ^2^ tests. Binary logistic regression model was primarily used to analyze the association between workplace cleaning agent exposures and adult asthma, while multinomial logistic regression was employed for asthma subtypes. Ever exposure to any cleaning agents (yes or no), exposure frequency (>weekly, yes or no), and duration (>1 year, yes or no) were examined as independent variables separately. Specific cleaning agents with high exposure proportions, including ammonia and chlorine bleach, were also analyzed. All models were adjusted for age, sex, the first five genetic PCs (or race in models without genetic factors), highest education, annual household income, employment status, smoking, and BMI. The missing data of covariates (0.3% - 2.2%) (**Table S1**) were singly imputed ^29^ using the random forest algorithm from the Multivariate Imputation by Chained Equations (Mice) package in R 4.2.0.

### G ⊆ E interaction analyses

#### (i) Development of polygenic risk scores (PRS)

ia) Genome-wide PRS: To ensure the generalization of our findings on genome-wide PRS, we used summary statistics of genome-wide association study (GWAS) of asthma from UK Biobank ^30^ and meta-analysis across 18 global biobanks. ^31^ The SNP weights for PRS were deposited in the [PGS Catalogue](https://www.genome.jp/pathway/map05310). After variant and allele harmonization process, seven candidate PRS were calculated in PEGS participants using PLINK 2.0 score function, as described previously. ^26^ The optimal PRS was derived from 68,835 individuals of White British ancestry ([Global Biobank Engine](https://biobankengine.stanford.edu/RIVAS_HG19/snpnet/BIN22127)) ^30^, which showed the maximal area under the curve (AUC) for asthma prediction in our samples (**Figure S2**). Age, sex, and the first five genetic PCs were adjusted.

ib) Pathway-based PRS: They were developed using a new feature called PRSet in PRSice v2.3.5 ^18^. *In prior* (**Figure S3)**, we identified curated gene sets for oxidative stress response (WikiPathways: WP408) and type 2 immune response (adapted from Gene Ontology biological process: 0042092 and Kyoto Encyclopedia of Genes and Genomes pathway: map05310). SNPs within 1 kilobase upstream and downstream of the 5’ and 3’ untranslated regions (UTRs) were annotated into targeted genes and analyzed for associations with asthma to get estimated weights. The best-fit PRS for each pathway was developed independently with the clumping and thresholding (C+T) method. Summary data for the best pathway-based PRS are presented in **Table S2,** and additional details and code can be found in the **supplementary methods** and https://github.com/Chenyj336/G-and-E-interaction_pathway-based-approach/.

#### (ii) Interaction analysis for genome-wide and pathway-based PRS

For multiplicative interactions, the product term of continuous genetic scores and cleaning agent exposure was induced in the logistic model, and multiplicative interaction *p* value was calculated. Meanwhile, PRS-related asthma risk was plotted for exposed and non-exposed groups to visualize effect modification by environmental factors. For additive interactions, PRS was dichotomized into low and high genetic risk groups based on the median value. The relative excess risk due to interaction (RERI) and proportion attributable to interaction (AP) were calculated, with values greater than zero indicating a positive additive interaction (i.e., joint excess risk *>* sum of individual excess risks) (**Supplementary methods**). Analyses were conducted using “*visreg”*and “*interactionR*” packages.

#### (iii) Gene-based interaction

To further explore key genes involved in pathway-based PRS interaction analysis, we used a multiple linear principal components regression model in MAGMA v1.10. This method applies an F-test to compute the *p*-values for joint interaction effects between the PCs of SNPs matrix of a gene and environmental factor ^32^. A Bonferroni corrected threshold of 0.05 (*P*=0.05/17 and 0.05/29) was used to identify significant interactive genes.

##### Sensitivity analyses

We developed pathway-based PRS based on candidate genes with 2- and 5-kilobase extended flanking regions, and re-analyzed their additive interaction with cleaning agents, respectively. We also conducted interaction analyses for different subtypes of asthma to explore potential heterogeneity in asthma pathogenesis.

## Results

### General characteristics of study population

**Table 1** summarizes the general characteristics of the 2615 individuals in this study, with a mean age of 50.90 years and 69.22 % females. There were 87.19% White and 9.22% Black individuals. Of the total, 250 adults (9.56%) reported current asthma, with 157 (6.00%) diagnosed in adulthood. Adults had current asthma were more likely to be female, Black or other minority races, obesity, and to have lower household incomes compared with those without asthma (*P* < 0.05).

**Table 1.** General characteristics of study population (n=2615)

|  | Total (N=2615) | Current asthma |  | p value |
| --- | --- | --- | --- | --- |
|  |  | No (n=2365) | Yes (n=250) |  |
| Age, year, Mean (SD) | 50.90 (14.18) | 50.98 (14.28) | 50.20 (13.22) | 0.409 |
| Sex, n (%) |  |  |  | **< 0.001** |
| Male | 805 (30.78) | 757 (32.01) | 48 (19.20) |  |
| Female | 1810 (69.22) | 1608 (67.99) | 202 (80.80) |  |
| Race, n (%) |  |  |  | **0.028** |
| White | 2280 (87.19) | 2075 (87.74) | 205 (82.00) |  |
| Black | 241 (9.22) | 207 (8.75) | 34 (13.60) |  |
| Others | 94 (3.59) | 83 (3.51) | 11 (4.40) |  |
| Highest education level, n (%) |  |  |  | 0.729 |
| High school or below | 121 (4.64) | 113 (4.79) | 8 (3.20) |  |
| College, technical or vocational school graduate | 576 (22.09) | 520 (22.06) | 56 (22.40) |  |
| Bachelor | 867 (33.26) | 783 (33.22) | 84 (33.60) |  |
| Graduate or professional | 1043 (40.01) | 941 (39.92) | 102 (40.80) |  |
| N-Miss | 8 | 8 | 0 |  |
| Annual household income, n (%) |  |  |  | **0.002** |
| Less than \$20,000 | 138 (5.39) | 112 (4.85) | 26 (10.44) | |
| \$20,000 to 49,999 | 547 (21.38) | 492 (21.31) | 55 (22.09) | |
| \$50,000 to 79,999 | 724 (28.30) | 657 (28.45) | 67 (26.91) | |
| \$80,000 or above | 1149 (44.92) | 1048 (45.39) | 101 (40.56) | |
| N-Miss | 57 | 56 | 1 |  |
| Employment, n (%) |  |  |  | 0.088 |
| No | 757 (28.95) | 673 (28.46) | 84 (33.60) |  |
| Yes | 1858 (71.05) | 1692 (71.54) | 166 (66.40) |  |
| BMI, n (%) |  |  |  | **< 0.001** |
| <25 kg/m<sup>2</sup> | 972 (37.44) | 905 (38.58) | 67 (26.80) |  |
| 25-<30 kg/m<sup>2</sup> (overweight) | 839 (32.32) | 772 (32.91) | 67 (26.80) |  |
| ≥30 kg/m<sup>2</sup> (obesity) | 785 (30.24) | 669 (28.52) | 116 (46.40) |  |
| N-Miss | 19 | 19 | 0 |  |
| Smoking status, n (%) |  |  |  | 0.456 |
| No | 1672 (64.11) | 1507 (63.88) | 165 (66.27) |  |
| Yes | 936 (35.89) | 852 (36.12) | 84 (33.73) |  |
| N-Miss | 7 | 6 | 1 |  |

Data presented were mean (SD) and n (%).
P value derived from two-sample t-test or $\chi^2$ -test

### Associations between cleaning agents, PRS, and adult asthma

**Table 2** shows that 25.8% (676/2615) of adults had ever been exposed to cleaning agents at work. The most reported cleaning agent was chlorine bleach (23.87%), followed by ammonia (8.18%). Ever exposure to any cleaning agent, as well as specific cleaning agents, was associated with increased odds of adult asthma in the adjusted model (OR = 1.56 ∼ 2.24). **Figure S4** illustrates the distribution of established PRS and their positive linear relationships with asthma.

**Table 2.** Association between occupational exposure to cleaning agents and asthma in adults (n=2615)

|  | Asthma case (n) /<br>Exposure (N)<sup>a</sup> | Crude model |  | Adjusted model |  |
| --- | --- | --- | --- | --- | --- |
|  |  | OR (95% CI) | *P* | OR (95% CI) | *P* |
| **Any cleaning agent** |  |  |  |  |  |
| Ever exposure | 91 / 676 | 1.74 (1.32, 2.29) | **<0.001** | 1.56 (1.18, 2.07) | **0.002** |
| Exposure frequency > weekly | 72 / 458 | 2.06 (1.53, 2.76) | **<0.001** | 1.82 (1.34, 2.47) | **<0.001** |
| Exposure duration>1 year | 83 / 610 | 1.76 (1.32, 2.33) | **<0.001** | 1.59 (1.19, 2.12) | **0.001** |
| **Specific cleaning agents** |  |  |  |  |  |
| Ever exposure to ammonia | 41 / 214 | 2.49 (1.70, 3.56) | **<0.001** | 2.24 (1.51, 3.25) | **<0.001** |
| Ever exposure to chlorine bleach | 88 / 624 | 1.85 (1.40, 2.44) | **<0.001** | 1.66 (1.24, 2.20) | **<0.001** |

<sup>a</sup> Asthma case number in participants with exposure
The adjusted logistic regression model included age, sex, race, employment, the highest education level, household income, smoking status, and BMI.

### Multiplicative interaction with PRS on adult asthma

Figure 2 illustrates the positive linear relationships between continuous PRS and asthma risk, modified by cleaning agent exposure. For genome-wide PRS, the positive associations with asthma were stronger in the groups exposed to any cleaning agent and bleach (*P* _interaction_ = 0.029 and 0.039), compared to non-exposure group. Pathway-based PRS for oxidative stress and type 2 immune response were positively associated with asthma in all groups, regardless of exposure status.

**Figure 2.**
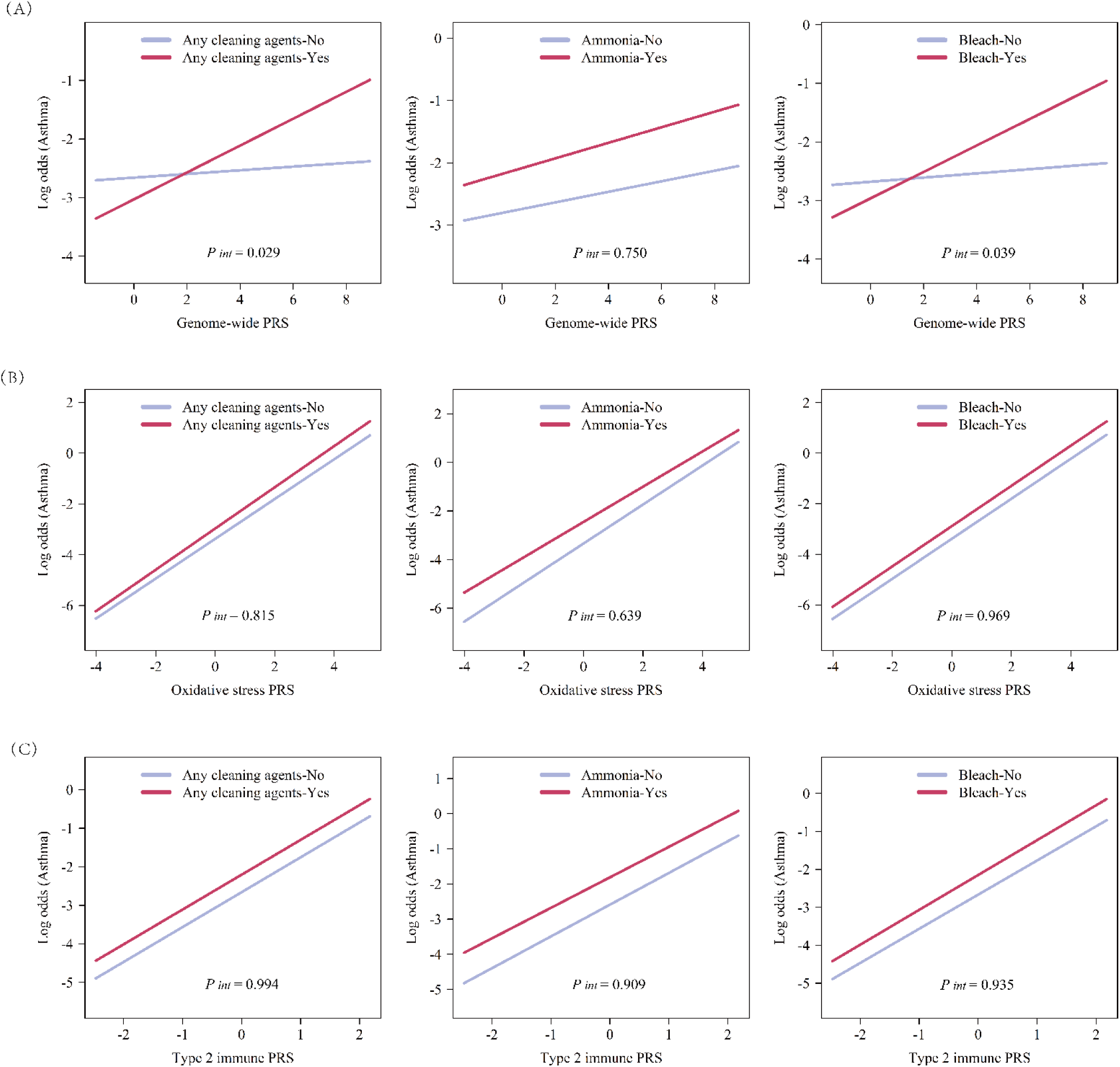
Association between continuous PRS and asthma modified by occupational cleaning agent exposure (n=2615) (A) Genome-wide PRS; (B) Oxidative stress pathway-based PRS; (C) Type 2 immune response-based PRS Abbreviations: PRS, polygenic risk score *P _int_*: Multiplicative interaction *P* value. The multiplicative interaction model was adjusted for age, sex, the first five genetic components, employment, the highest education level, household income, smoking, and BMI.

### Additive interaction with PRS on adult asthma

Figure 3 and Figure 4 show the additive interactions of PRS-based genetic risk and cleaning agent exposure on asthma. Asthma risk was strongest in the group with both high genetic risk and cleaning agent exposure, compared to the group with low genetic risk and no exposure.

**Figure 3.**
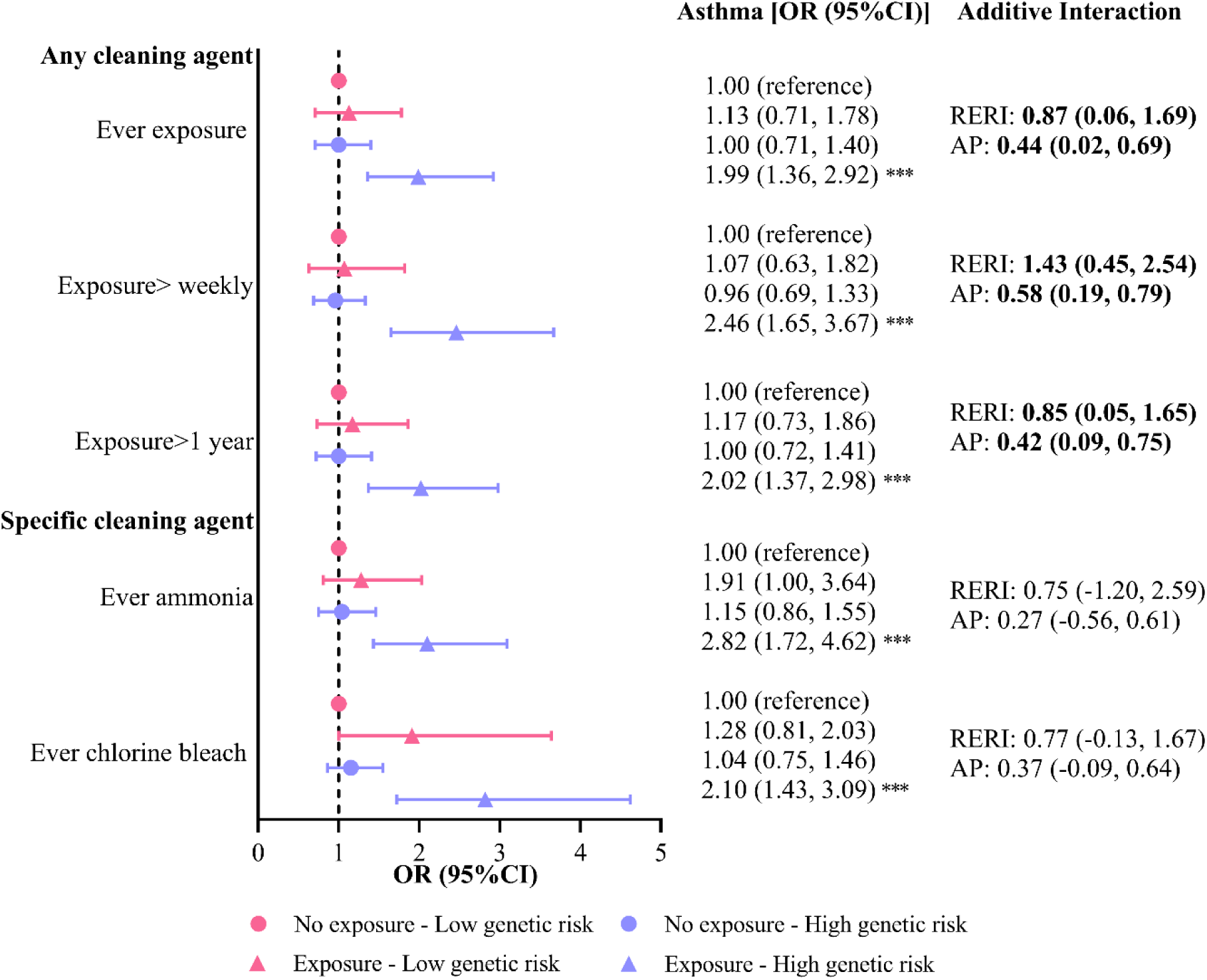
Additive interaction of occupational cleaning agents and genome-wide PRS on adult asthma (n=2615) Abbreviation: RERI, relative excess risk due to interaction (part of the total effect due to interaction); AP, proportion attributable to interaction (proportion of the combined effect due to interaction). The PRS was dichotomized into high and low genetic risk groups based on the median value. The logistic regression model was adjusted for age, sex, the first five genetic components, employment, the highest education level, household income, smoking status, and BMI. A RERI or AP greater than zero indicated significant additive interaction (in bold). * *P* <0.05; \*\**P*<0.01; \*\*\**P*<0.001.

**Figure 4.**
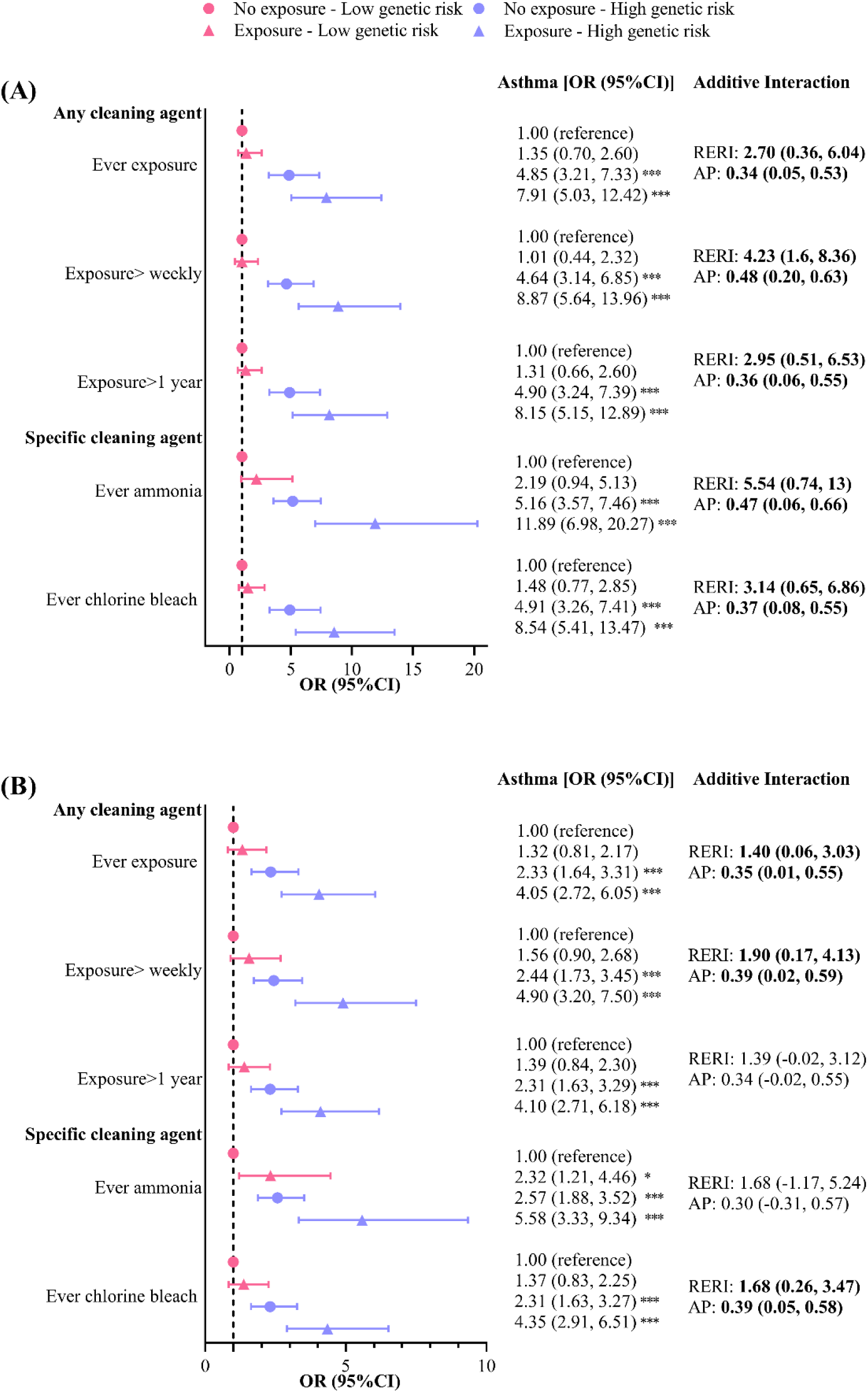
Additive interaction of (A) oxidative stress and (B) type 2 immune response pathway-based PRS and occupational cleaning agents on adult asthma (n=2615) Abbreviation: RERI, relative excess risk due to interaction (part of the total effect due to interaction); AP, proportion attributable to interaction (proportion of the combined effect due to interaction). The PRS was dichotomized into high and low genetic risk groups based on the median value. The logistic regression model was adjusted for age, sex, the first five genetic components, employment, the highest education level, household income, smoking status, and BMI. A RERI or AP greater than zero indicated significant additive interaction (in bold). * *P* <0.05; \*\**P*<0.01; \*\*\**P*<0.001.

Specifically, exposure to any cleaning agent additively interacted with high genome-wide risk, suggesting a 0.87 relative excessive risk due to interaction, accounting for 44% of their joint effect (OR=1.99, 95% CI: 1.36, 2.92) in Figure 3. For pathway-based PRS (Figure 4), we consistently found positive additive interactions between any or specific cleaning agents and oxidative stress pathway-related risk (OR=7.91-11.89 for joint effect). Type 2 immune pathway-related PRS also synergistically interacted with any cleaning agents and bleach (OR = 4.05-5.58 for joint effect).

### Gene-based interaction on adult asthma

As shown in **Tables S3** and **S4**, significant interactions were observed between exposure to any or specific cleaning agents and genes in the oxidative stress pathway, including *XDH*, *UGT1A6*, *MAPK10*, *MAPK14*, *GSR*, *GCLC*, *SP1*, and *NOX5* (*P* _interaction_ < 0.05/29 genes; 1.72⊆10^-3^). Suggestive, but not statistically significant, interactions were found for genes in the type 2 immune response pathway, including *BCL6*, and *MYB* (*P* _interaction_ < 0.10/17 genes; 5.88⊆10^-3^).

### Sensitivity analysis

In **Figures S5**-**S7**, consistent additive interactions were found for pathway-based PRS when extending 2 and 5 kilobase upstream and downstream gene regions. When analyzed the subtypes of adult asthma, we found genetic liability to oxidative stress risk specifically interacted with cleaning agents on adult-exacerbated asthma (childhood-onset), especially for weekly exposure to cleaning agents (**Table S6**). Genome-wide risk showed interactive effects with any cleaning agent exposure for adult-onset asthma (**Table S7**).

## Discussion

This study demonstrated an increased risk of current asthma in adults associated with workplace exposure to cleaning agents, including ammonia and bleach. Genome-wide PRS predisposed adults to a higher asthma risk when exposed to any cleaning agent. We also found additive interaction between cleaning agent exposure and genetic risks for asthma, particularly involving oxidative stress and type 2 immune response pathways. Gene-based analysis further suggested potential interactive roles of stress-regulating and antioxidant genes in the pathway. These findings provide mechanistic insights into occupational asthmagens and underscore the importance of targeted intervention strategies for susceptible populations.

Exposure to cleaning products has been reported as one of the most prevalent causes of work-related asthma. ^4^ Several occupational cohorts among cleaning workers and nurses have identified cleaning agents as risk factors for new-onset asthma and poorly controlled symptoms, especially among females. ^33–35^ Our results supported previous findings, showing similar positive associations in the population with substantial cleaning agent exposure (25.8%) and a predominance of women (69.2%). As cleaning products are complex chemical mixtures, identifying the causative components for asthma risk remains challenging. Nonetheless, our findings suggest that bleach (i.e., chlorine and hypochlorite) and ammonia may play crucial roles. Bleaching agents and ammonia are known irritants that can cause acute airway injury, and they also contribute to the formation of various hazardous pollutants, including VOCs (e.g., chloroform and formaldehyde), organic particulate matter, and nitrogen dioxide, all of which can adversely impair respiratory health. ^19,36,37^ However, the exact pathophysiological mechanisms by which cleaning agents influence asthma risk and the modulating effects of genetic factors are poorly known.

GWAS have highlighted the polygenic nature of asthma. ^38^ Two previous genome-wide interaction studies have scanned millions of SNPs to identify novel loci interacting with occupational exposures on respiratory symptoms or lung function. ^21,22^ However, these studies did not specifically examine cleaning products, and such data-driven strategies require large sample sizes for effective discovery. Our study employed a more statistically powerful tool—PRS—revealing the association of genome-wide PRS with asthma was modified by cleaning agent exposure. We estimated genome-wide PRS using summary statistics from UK Biobank (British ancestry) ^30^, as it showed better predictive performance in our multi-ancestry population (87.2% White and 9.2% Black individuals) compared with PRS derived from a global biobank meta-analysis. ^31^ Genome-wide approach improves generalization of the observed G ⊆ E interaction for heterogenous asthma, which aggregated the risk of genetic variants across distinct pathogeneses, including those with small or moderate effects. On the other hand, pathway-based PRS integrates the advantages of polygenic scores and pathway-specific strategies. The predefined pathways showed positive associations with asthma in groups exposed and unexposed to cleaning agents in this study, suggesting their effects cannot be simply multiplicatively decomposed but involve interactions with broader exogenous or endogenous factors of asthma.

Pathway-based additive interaction analysis provides a biologically interpretable perspective, and we found cleaning agents synergistically interacted with oxidative stress-related genetic risk for asthma. Oxidative stress results from an imbalance between the production of reactive oxygen species and the capacity of the antioxidant defense mechanism. Bleaching agents are oxidants with high positive electrochemical standard potential, while ammonia inhalation has been shown to decrease antioxidant enzymes. ^40^ These toxicants may induce oxidative stress and epithelium injury by interacting with genetic susceptibility, leading to asthma initiation, hyperreactivity, and remodeling ^12^ To our knowledge, only one study has employed the pathway-based selection process to analyze how occupational irritants interact with genetic predispositions to oxidative stress, identifying genes related to the nuclear factor-ƘB pathway. ^20^ We found similar interactive effects using pathway-based PRS, while gene-based analysis reveals that the interaction may be partly attributable to genes related to the mitogen-activated protein kinase (MAPK) pathway (e.g., *MAPK10* and *MAPK14*) and antioxidant enzymes in glutathione synthesis (e.g., *GSR*, and *GCLC*). The MAPK pathway responds to high levels of oxidative stimuli and can further activate transcription factors and pro-inflammatory intracellular signaling cascades. ^41^ It is also thought to activate the *AP1* family, which regulates antioxidant genes and asthma-associated cytokines. ^42^ Meanwhile, genetic variation in the glutathione synthesis pathway has been associated with differences in susceptibility to environmental hazards on lung function. ^14^ These findings provide insights for further studies to elucidate the transcriptional regulation and signaling pathways of oxidative stress in asthma.

We also found that type 2 immune response-related genetic risk interacted with any cleaning agent, suggesting T helper 2 (Th2) cell-mediated allergic inflammation plays a role in occupational asthma. While the irritant property of cleaning agents is more commonly recognized, potential sensitizers, such as quaternary ammonium compounds, chlorhexidine, and glutaraldehyde, have been reported. ^3,43,44^ These antigens might activate mast cells and Th2 cells, leading to the production of inflammatory cytokines (e.g., interleukin-4, −5, and −13) and mediators (e.g., leukotrienes and granule proteins), triggering an IgE-mediated immune response ^11^. However, our study did not identify specific cleaning sensitizers, and instead highlighted the interactive effects of irritant agents like chlorine bleach. Nonetheless, recent research has suggested that cleaning irritants may increase allergen permeability, possibly by disrupting epithelial barriers, which could initiate skin sensitization and lead to delayed-type hypersensitivity from airborne cleaning chemicals. ^45^ Exposure to chlorine has been shown in animal studies to potentiate Th2 responses and exacerbate airway allergic inflammation. ^46,47^ Therefore, the observed interactive effects between “any cleaning agent” and genetic risk of Th2 immune response may result from a mixture of exposure to irritants and sensitizers. However, gene-based interactions did not reach statistical significance after Bonferroni correction within this pathway, indicating that the underlying immunological mechanisms warrant further detailed exploration.

This study has several strengths. Most gene-by-exposure studies tested multiplicative interaction to imply effect modifications of genetic variants due to exposure, or vice versa. ^39^ Our study novelly analyzed pathway-based PRS to reveal the additive G ⊆ E interaction, supporting pathway-based PRS as an effective tool to uncover the biological depths of complex diseases and serve as a semi-explorative approach to help target key functional genes. ^48^ Our results were robust when including upstream and downstream correlated genetic variants near candidate genes in selected pathways. We also differentiated asthma subtypes by age of diagnosis and consistently found that genetic risk interacted with cleaning agent-induced asthma risk. While age-of-onset subtypes may share common genetic architectures ^38^, our result suggested that the oxidative stress pathway plays a more important role in exacerbating childhood-onset asthma during adulthood.

Several limitations of this study are acknowledged. First, asthma and cleaning exposure were self-reported using standardized questionnaires, limiting our ability to verify causal chemicals and confirm clinical plausibility. Given that cleaning products are complex mixtures, previous studies have largely relied on questionnaire to investigate information on exposure frequency, duration, and common chemical agents. ^49^ Second, we only focused on curated gene sets based on two well-established biological pathways for asthma. However, hypothesis-free strategies may uncover novel mechanisms in future studies with large sample sizes. Third, while this study primarily used PRS to summarize genetic risks, we did not examine interactions at the specific SNP level. Future research should investigate gene variants of candidate genes suggested to interact with cleaning agents in our study. Fourth, due to the cross-sectional design, we could not establish temporal relationships, nor rule out unmeasured confounding residuals (e.g., the use of household cleaning products and other occupational hazards). Nevertheless, we adjusted for smoking and obesity in the analysis, which are known sources of oxidative stress. Fifth, although our participants were recruited from the general population, the sample was predominantly White and female. Our findings should be further validated in other racial groups and caution should be exercised when generalizing to other populations.

## Conclusion

This study is the first to reveal biological interactions between occupational cleaning agents and genetic susceptibility to asthma from multi-level perspectives. Genome-wide PRS comprehensively evaluated aggregated genetic risks, while pathway-based PRS provided interpretable mechanistic insights for G⊆E interactions on asthma. These findings help to develop precision prevention and intervention strategies for asthma, particularly in individuals with genetic susceptibility related to oxidative stress and type 2 immune pathways.

## Data Availability

The data will be available under controlled access to comply with polices and process of the PEGS data and meet applicable NIH requirements for data sharing and that the privacy and confidentiality of human subjects are protected. Researchers seeking access to PEGS data may contact the PEGS Executive Leadership Committee with details from the PEGS websites

