## Supplementary methods for "Pathway-based genetic susceptibility and cleaning agent exposures in adult asthma: A semi explorative GxE analysis in the Personalized Environment and Gene Study (PEGS)"

**Supplementary M****ethods**

**Study population**

**Figure S1** illustrates the details of sample exclusion and quality control of WGS data to achieve final samples for PRS development and further analysis. Additional details about the study design and questionnaires can be found at <https://www.niehs.nih.gov/research/clinical/studies/pegs/index.cfm>.

**PRS development**

**(1) Genome-wide PRS.** The meta data of finally selected genome-wide PRS for doctor-diagnosed asthma can be found in the PGS Catalog (<https://www.pgscatalog.org/score/PGS001345/>) and [Global Biobank Engine](https://biobankengine.stanford.edu/RIVAS_HG19/snpnet/BIN22127) ([Tanigawa et al., PLOS Genet. 2022](10.1371/journal.pgen.1010105)). It was developed based on the model development set of 68,835 white British ancestry in UK Biobank data. The predictive performance was evaluated on a hold-out test set of 15,031 white British ancestry. The transferability of the model was evaluated on the additional sets of individuals of other ancestry groups, including non-British white, south Asian, East Asian, and African. The predictive performance of genome-wide PRS in PEGS participants is shown in Figure S2.

**(2) Pathway-based PRS.** GWAS for the candidate pathways was conducted using PLINK 1.9 logistic function. The summary statistics for the final pathway-based PRS are showed in the **supplementary Excel sheets**. Optimal PRS were developed with PRSice v2.3.5. Genome boundary file was download from Ensembl browser (Ch38.112.gtf); MSigDB file was created based on the gene in two hypothesized pathways. We used “--clump-kb 1M --clump-r^2^ 0.1” for LD clumping and “--lower 0.01 --upper 1” for *P*-value thresholding. Since SNPs outside the gene region are also likely to influence functions of the gene, we added one kilobase to the 3' region of each gene regions and included pairwise SNPs with LD>0.8 within the regions (--wind-3 1k --wind-5 –1k --proxy 0.8) in the main analysis, and then tested 2 and 5 kb in the sensitivity analyses. We used direct summation of the effect size “--score sum” to calculate PRS. Age, sex, and the first five genetic PCs were adjusted. The R^2^ of PRS and the best P threshold was shown in **Table S1.**

**(3) Addictive interaction indexes**

RERI and AP were calculated according to the following formula [1] and [2], respectively.

$RERI={OR}_{G+E+}-{OR}_{G+E-}-{OR}_{G-E+}+1$ ………………………………………………[1]

$AP=\frac{RERI}{{OR}_{G+E+}}$ ………………………………………………………………………………[2]

For two dichotomous factors G (dichotomous PRS) and E (cleaning agent exposure): OR_G+E+_ is the relative risk of disease if both G and E are present, OR_G+E−_ is the relative risk of disease if G is present but factor E is absent, OR_G-E+_ is the relative risk of disease if G is absent but E is present.

**Gene-based interaction in the hypothesized pathways**

We used MAGMA (<https://ctg.cncr.nl/software/magma>) to project the SNP matrix for a gene onto its PCs with raw genotype data and then induces pruned PCs (>99.9% of the variance) and the interactor (environment exposure) for asthma in the linear principal component regression model. To include SNPs in a window around genes, a 1 kilobase symmetrical annotation window was added in 3’ end and 5’ end (--annotate window=1). A gene by environmental factor interaction component was included in the regression model, and p value for the joint interaction effects was computed. The method was reported to have greater statistical power for gene-based analysis..

**Supplementary Tables and Figures**

Table S1 The distribution of variables in complete and imputed data sets (N=2615)

|  | Original data set | Imputed data set | *P* value *^a^* |
| --- | --- | --- | --- |
| **BMI, n (%)** |  |  | 0.988 |
| <25 kg/m^2^ | 972 (37.4) | 983 (37.6) |  |
| 25-<30 kg/m^2^ (overweight) | 839 (32.3) | 840 (32.1) |  |
| ≥30 kg/m^2^ (obesity) | 785 (30.2) | 792 (30.3) |  |
| N-Miss | 19 | 0 |  |
| **Smoking status, n (%)** |  |  | 0.989 |
| No | 1672 (64.1) | 1676 (64.1) |  |
| Yes | 936 (35.9) | 939 (35.9) |  |
| N-Miss | 7 | 0 |  |
| **Highest education level, n (%)** |  |  | 1.000 |
| High school or below | 121 (4.6) | 121 (4.6) |  |
| College, technical or vocational school graduate | 576 (22.1) | 578 (22.1) |  |
| Bachelor | 867 (33.3) | 871 (33.3) |  |
| Graduate or professional | 1043 (40.0) | 1045 (40.0) |  |
| N-Miss | 8 | 0 |  |
| **Annual household income, n (%)** |  |  | 0.962 |
| Less than $20,000 | 138 (5.4) | 150 (5.7) |  |
| $20,000 to 49,999 | 547 (21.4) | 556 (21.3) |  |
| $50,000 to 79,999 | 724 (28.3) | 736 (28.1) |  |
| $80,000 or above | 1149 (44.9) | 1173 (44.9) |  |
| N-Miss | 57 | 0 |  |

Data presented were n (%).

*^a^ P* value derived from χ^2^-test to compare the difference between original and imputed data sets

Table S2 The summary statistics for the best pathway-based PRS

| Pathway | PRS R^2^ | β | SE | *P* | SNP number |
| --- | --- | --- | --- | --- | --- |
| Oxidative stress response | 0.184 | 0.781 | 0.056 | 9.85E-44 | 193 |
| Type 2 immune response | 0.063 | 0.906 | 0.106 | 1.29E-17 | 55 |

The GWAS summary statistics for pathway-based PRS are showed in the **supplementary Excel sheets**

Table S3 Gene-based interaction in the oxidative stress pathway with occupational exposure on asthma in the linear principal component regression

| Gene | CHR | Position | | SNP number | P for interaction with exposure | | | | |
| --- | --- | --- | --- | --- | --- | --- | --- | --- | --- |
|  |  | Start | Stop |  | Any cleaning agent | Cleaning agent -  frequency | Cleaning agent -duration | Ammonia | Bleach |
| XDH | 2 | 31333321 | 31415742 | 146 | 1.22E-01 | 5.15E-03 | 1.43E-01 | **2.51E-04** | 1.49E-01 |
| NFE2L2 | 2 | 177217667 | 177393756 | 37 | 4.94E-01 | 6.36E-02 | 7.23E-01 | 5.69E-01 | 5.40E-01 |
| UGT1A6 | 2 | 233690607 | 233774300 | 203 | 2.03E-03 | **1.63E-03** | 5.01E-02 | 1.45E-02 | **6.13E-04** |
| GPX1 | 3 | 49356174 | 49359605 | 4 | 5.74E-01 | 4.82E-01 | 7.81E-01 | 1.85E-01 | 5.29E-01 |
| SOD3 | 4 | 24788912 | 24801842 | 5 | 2.53E-01 | 3.76E-01 | 2.16E-01 | 1.68E-01 | 5.18E-01 |
| MAPK10 | 4 | 85989007 | 86595625 | 853 | 3.13E-02 | 4.22E-02 | 6.20E-02 | **5.86E-05** | 7.29E-02 |
| NFKB1 | 4 | 102500330 | 102618302 | 152 | 7.73E-01 | 4.80E-01 | 7.52E-01 | 1.87E-01 | 8.46E-01 |
| GPX3 | 5 | 151019591 | 151029988 | 33 | 4.28E-01 | 5.12E-01 | 5.63E-01 | 3.74E-01 | 4.47E-01 |
| MAPK14 | 6 | 36026782 | 36112236 | 150 | 5.51E-02 | **1.42E-03** | 7.86E-02 | 4.11E-02 | 8.71E-02 |
| GCLC | 6 | 53496341 | 53617970 | 86 | 8.01E-01 | 3.15E-01 | 7.54E-01 | **8.05E-04** | 5.41E-01 |
| NOX3 | 6 | 155394368 | 155456839 | 108 | 3.47E-01 | 1.89E-01 | 2.14E-01 | 2.72E-03 | 4.10E-01 |
| SOD2 | 6 | 159668069 | 159763529 | 195 | 9.70E-02 | 4.41E-03 | 4.00E-02 | 2.02E-01 | 9.62E-02 |
| GSR | 8 | 30677066 | 30728846 | 54 | 6.52E-03 | **1.42E-03** | 6.03E-03 | 9.49E-03 | 3.58E-03 |
| CAT | 11 | 34437934 | 34473060 | 73 | 3.16E-01 | 3.40E-01 | 8.70E-01 | 4.24E-02 | 5.36E-01 |
| NOX4 | 11 | 89323353 | 89499187 | 177 | 9.12E-02 | 6.85E-02 | 8.98E-02 | 1.48E-01 | 2.27E-01 |
| MGST1 | 12 | 16346142 | 16610259 | 322 | 4.56E-01 | 3.59E-01 | 3.11E-01 | 5.28E-03 | 5.44E-01 |
| SP1 | 12 | 53379176 | 53417446 | 44 | 3.01E-03 | **2.60E-06** | 4.35E-03 | 4.36E-02 | **9.41E-04** |
| TXNRD1 | 12 | 104214779 | 104351307 | 341 | 9.28E-02 | 2.78E-01 | 1.42E-01 | 2.29E-02 | 4.99E-02 |
| FOS | 14 | 75277826 | 75283230 | 7 | 6.97E-01 | 2.11E-01 | 8.22E-01 | 1.28E-01 | 5.36E-01 |
| NOX5 | 15 | 69013695 | 69063762 | 17 | 3.27E-01 | 1.40E-01 | 4.36E-01 | **1.21E-03** | 4.01E-01 |
| CYP1A1 | 15 | 74718542 | 74726536 | 2 | 2.18E-01 | 2.91E-01 | 5.01E-01 | 3.16E-02 | 4.82E-01 |
| MT1X | 16 | 56681470 | 56685196 | 10 | 1.57E-01 | 1.09E-01 | 3.25E-01 | 2.98E-02 | 3.06E-01 |
| NQO1 | 16 | 69705996 | 69727668 | 30 | 2.47E-01 | 1.86E-01 | 3.76E-01 | 1.91E-01 | 2.06E-01 |
| JUNB | 19 | 12790486 | 12794315 | 2 | 2.33E-02 | 4.46E-02 | 4.92E-02 | 1.49E-01 | 9.14E-03 |
| NFIX | 19 | 12994475 | 13099796 | 52 | 8.11E-01 | 3.47E-01 | 6.85E-01 | 9.26E-01 | 8.79E-01 |
| SOD1 | 21 | 31658666 | 31669931 | 7 | 3.55E-01 | 5.72E-02 | 3.19E-01 | 1.78E-01 | 2.45E-01 |
| TXNRD2 | 22 | 19874517 | 19942820 | 181 | 3.13E-01 | 6.26E-02 | 2.01E-01 | 2.03E-01 | 1.96E-01 |
| HMOX1 | 22 | 35379361 | 35395214 | 19 | 4.80E-02 | 4.07E-03 | 7.02E-02 | 5.86E-01 | 6.37E-02 |
| TXN2 | 22 | 36466046 | 36482640 | 49 | 8.97E-01 | 3.78E-01 | 9.54E-01 | 7.96E-01 | 9.52E-01 |

Bonferroni corrected P=0.05/29=1.72E-03; 0.10/29=3.45E-03

Extended 1kb base-pair upstream and downstream of coding area

The logistic regression model was adjusted for age, sex, the first five genetic PCs, employment, the highest education level, household income, smoking status, and BMI

Table S4 Gene-based interaction in the type 2 immune pathway with occupational exposures on asthma in the linear principal component regression

| Gene | CHR | Position | | SNP number | P for interaction with exposure | | | | |
| --- | --- | --- | --- | --- | --- | --- | --- | --- | --- |
|  |  | Start | Stop |  | Any cleaning agent | Cleaning agent  frequency | Cleaning agent Duration | Ammonia | Bleach |
| FCER1A | 1 | 159288714 | 159309224 | 9 | 3.47E-01 | 7.90E-01 | 1.91E-01 | 1.13E-01 | 6.07E-01 |
| IL10 | 1 | 206766602 | 206775541 | 17 | 5.56E-01 | 2.27E-01 | 6.14E-01 | 5.74E-01 | 7.41E-01 |
| HLX | 1 | 220878431 | 220886059 | 18 | 3.14E-01 | 2.93E-01 | 4.40E-01 | 4.12E-01 | 3.37E-01 |
| BCL6 | 3 | 187720377 | 187746725 | 36 | 6.82E-01 | 2.61E-01 | 7.51E-01 | 4.04E-03 | 7.81E-01 |
| IL5 | 5 | 132540445 | 132557838 | 4 | 9.36E-01 | 9.88E-01 | 4.82E-01 | 3.45E-01 | 8.35E-01 |
| IL13 | 5 | 132655263 | 132662110 | 8 | 9.58E-01 | 5.95E-01 | 9.24E-01 | 9.38E-01 | 9.57E-01 |
| IL4 | 5 | 132672986 | 132683678 | 3 | 4.22E-01 | 2.45E-01 | 5.00E-01 | 4.39E-01 | 3.79E-01 |
| TRAF3IP2 | 6 | 111554381 | 111607906 | 99 | 2.70E-01 | 3.91E-01 | 3.99E-01 | 2.79E-01 | 1.59E-01 |
| MYB | 6 | 135180308 | 135220173 | 43 | 2.01E-02 | 1.28E-02 | 1.60E-01 | 5.63E-03 | 7.34E-02 |
| IL33 | 9 | 6214786 | 6258983 | 81 | 1.93E-02 | 1.22E-01 | 2.68E-02 | 1.54E-01 | 1.10E-02 |
| GATA3 | 10 | 8044378 | 8076198 | 46 | 4.52E-01 | 1.13E-01 | 4.49E-01 | 9.55E-03 | 3.32E-01 |
| MEN1 | 11 | 64802510 | 64812294 | 4 | 2.81E-01 | 3.54E-01 | 3.60E-01 | 3.00E-01 | 3.50E-01 |
| IL18 | 11 | 112142253 | 112165096 | 36 | 8.28E-01 | 4.33E-01 | 6.85E-01 | 5.27E-01 | 9.12E-01 |
| KMT2A | 11 | 118435456 | 118527832 | 32 | 5.00E-01 | 6.73E-01 | 6.85E-01 | 4.57E-02 | 3.48E-01 |
| BATF | 14 | 75521455 | 75547993 | 36 | 2.52E-01 | 2.51E-01 | 1.39E-01 | 7.26E-01 | 1.73E-01 |
| IL4R | 16 | 27312668 | 27365778 | 96 | 8.90E-01 | 6.09E-01 | 8.16E-01 | 9.11E-02 | 8.34E-01 |
| BCL3 | 19 | 44746705 | 44761044 | 10 | 1.80E-01 | 2.47E-01 | 2.63E-01 | 9.25E-01 | 1.94E-01 |

Bonferroni corrected P=0.05/17=2.94E-03; 0.1/17=5.88E-03

Extended 1 kilobase upstream and downstream of coding area

The logistic regression model was adjusted for age, sex, the first five genetic PCs, employment, the highest education level, household income, smoking status, and BMI.

Table S5 Association between occupational exposure to cleaning agents and asthma subtypes by onset age (n=2615)

|  | Childhood-onset asthma | | Adult-onset asthma | |
| --- | --- | --- | --- | --- |
|  | OR (95% CI) | *P* | OR (95% CI) | *P* |
| **Any cleaning agents** |  |  |  |  |
| Ever exposure | 1.66 (1.06, 2.58) | 0.025 | 1.52 (1.07, 2.16) | 0.019 |
| Exposure frequency > weekly | 1.60 (1.01, 2.55) | 0.047 | 1.60 (1.12, 2.28) | 0.010 |
| Exposure duration>1 year | 2.05 (1.28, 3.28) | 0.003 | 1.72 (1.17, 2.52) | 0.005 |
| **Individual cleaning agent** |  |  |  |  |
| Ever exposure to ammonia | 2.91 (1.65, 5.11) | <0.001 | 1.89 (1.17, 3.07) | 0.009 |
| Ever exposure to chlorine bleach | 1.85 (1.19, 2.88) | 0.007 | 1.57 (1.10, 2.24) | 0.013 |

The multinomial logistic regression model was adjusted for age, sex, race, employment, the highest education level, household income, smoking status, and BMI.

Table S6 Interaction between occupational cleaning agent exposure and PRS on childhood-onset asthma (n=2458)

|  | Childhood-onset (adult-exacerbated) asthma ^a^ [OR (95% CI)] | | | | |
| --- | --- | --- | --- | --- | --- |
|  | Genome-wide risk |  | Oxidative stress pathway-related risk |  | Type 2 immune response-related risk |
| **Any cleaning agent** |  |  |  |  |  |
| **Ever exposure** |  |  |  |  |  |
| No exposure - Low G | 1.00 (reference) |  | 1.00 (reference) |  | 1.00 (reference) |
| No exposure - High G | 0.91 (0.53, 1.55) |  | 3.31 (1.82, 6.03) ^***^ |  | 2.36 (1.34, 4.13) ^**^ |
| Exposure - Low G | 1.25 (0.62, 2.50) |  | 0.83 (0.27, 2.53) |  | 1.42 (0.65, 3.10) |
| Exposure - High G | 1.87 (1.03, 3.39) ^*^ |  | 6.27 (3.27, 12.02) ^***^ |  | 4.27 (2.27, 8.03) ^***^ |
| Multiplicative *P* value | 0.278 |  | 0.185 |  | 0.612 |
| RERI (95% CI) | 0.71 (-0.72, 2.06) |  | **3.13 (0.29, 7.84)** |  | 1.5 (-0.90, 4.59) |
| AP (95% CI)) | 0.38 (-0.47, 0.80) |  | **0.50 (0.05, 0.72)** |  | 0.35 (-0.26, 0.64) |
| **Exposure> weekly** |  |  |  |  |  |
| No exposure - Low G | 1.00 (reference) |  | 1.00 (reference) |  | 1.00 (reference) |
| No exposure - High G | 0.87 (0.52, 1.48) |  | 3.02 (1.70, 5.36) ^***^ |  | 2.43 (1.39, 4.25) ^**^ |
| Exposure - Low G | 1.35 (0.62, 2.91) |  | 0.31 (0.04, 2.35) |  | 1.71 (0.73, 3.99) |
| Exposure - High G | 2.41 (1.30, 4.48) ^**^ |  | 7.33 (3.84, 13.97) ^***^ |  | 5.61 (2.88, 10.91) ^***^ |
| Multiplicative *P* value | 0.151 |  | **0.054** |  | 0.562 |
| RERI (95% CI) | 1.19 (-0.56, 3.09) |  | **5.00 (1.72, 10.85)** |  | 2.47 (-0.58, 7.06) |
| AP (95% CI)) | 0.49 (-0.32, 0.84) |  | **0.68 (0.28, 0.83)** |  | 0.44 (-0.16, 0.69) |
| **Exposure>1 year** |  |  |  |  |  |
| No exposure - Low G | 1.00 (reference) |  | 1.00 (reference) |  | 1.00 (reference) |
| No exposure - High G | 0.92 (0.54, 1.57) |  | 3.36 (1.85, 6.13) ^***^ |  | 2.36 (1.34, 4.13) ^**^ |
| Exposure - Low G | 1.29 (0.63, 2.65) |  | 0.71 (0.20, 2.48) |  | 1.40 (0.62, 3.15) |
| Exposure - High G | 1.76 (0.94, 3.29) |  | 6.26 (3.20, 12.21) ^***^ |  | 4.20 (2.18, 8.12) ^***^ |
| Multiplicative *P* value | 0.419 |  | 0.163 |  | 0.630 |
| RERI (95% CI) | 0.54 (-0.69, 1.77) |  | **3.18 (0.02, 6.35)** |  | 1.45 (-0.89, 3.78) |
| AP (95% CI)) | 0.31 (-0.32, 0.94) |  | **0.51 (0.20, 0.81)** |  | 0.34 (-0.09, 0.78) |
| **Specific cleaning agents** |  |  |  |  |  |
| **Ammonia** |  |  |  |  |  |
| No exposure - Low G | 1.00 (reference) |  | 1.00 (reference) |  | 1.00 (reference) |
| No exposure - High G | 1.03 (0.64, 1.66) |  | 3.58 (2.09, 6.16) ^***^ |  | 2.49 (1.51, 4.10) ^***^ |
| Exposure - Low G | 2.60 (1.04, 6.55) |  | 0.80 (0.10, 6.09) |  | 2.51 (0.92, 6.88) |
| Exposure - High G | 3.08 (1.47, 6.44) ^**^ |  | 11.63 (5.57, 24.26) ^***^ |  | 7.58 (3.55, 16.17) ^***^ |
| Multiplicative *P* value | 0.816 |  | 0.195 |  | 0.756 |
| RERI (95% CI) | 0.44 (-3.66, 3.82) |  | **8.25 (1.21, 20.03)** |  | 3.58 (-1.79, 11.63) |
| AP (95% CI)) | 0.14 (-1.59, 0.66) |  | **0.71 (0.14, 0.84)** |  | 0.47 (-0.38, 0.73) |
| **Bleach** |  |  |  |  |  |
| No exposure - Low G | 1.00 (reference) |  | 1.00 (reference) |  | 1.00 (reference) |
| No exposure - High G | 0.93 (0.54, 1.58) |  | 3.27 (1.80, 5.96) ^***^ |  | 2.33 (1.33, 4.09) ^**^ |
| Exposure - Low G | 1.44 (0.72, 2.88) |  | 0.90 (0.29, 2.75) |  | 1.54 (0.70, 3.37) |
| Exposure - High G | 2.04 (1.12, 3.71) ^*^ |  | 7.00 (3.65, 13.46) ^***^ |  | 4.78 (2.54, 9.00) ^***^ |
| Multiplicative *P* value | 0.355 |  | 0.164 |  | 0.555 |
| RERI (95% CI) | 0.68 (-0.93, 2.16) |  | **3.83 (0.84, 9.22)** |  | 1.91 (-0.62, 5.43) |
| AP (95% CI)) | 0.33 (-0.55, 0.74) |  | **0.55 (0.13, 0.74)** |  | 0.40 (-0.18, 0.66) |

a There were 93 individuals with asthma onset in childhood, and cases with adult-onset asthma were excluded in the analysis.

Abbreviation: G: genetic risk measured by PRS. RERI, relative excess risk due to interaction (part of the total effect due to interaction); AP, proportion attributable to interaction (proportion of the combined effect due to interaction).

The PRS was dichotomized into high and low genetic risk groups based on the median value.

The logistic regression model was adjusted for age, sex, the first five genetic components, employment, the highest education level, household income, smoking status, and BMI.

A RERI or AP greater than zero indicated significant additive interaction (in bold).

^*^ *P* <0.05; ^**^*P*<0.01; ^***^*P*<0.001.

Table S7 Interaction between occupational cleaning agent exposure and PRS on adult-onset asthma (n=2522)

|  | Adult-onset asthma ^a^ [OR (95% CI)] | | | | |
| --- | --- | --- | --- | --- | --- |
|  | Genome-wide risk |  | Oxidative stress pathway-related risk |  | Type 2 Immune response-related risk |
| **Any cleaning agent** |  |  |  |  |  |
| **Ever exposure** |  |  |  |  |  |
| No exposure - Low G | 1.00 (reference) |  | 1.00 (reference) |  | 1.00 (reference) |
| No exposure - High G | 1.04 (0.68, 1.58) |  | 6.31 (3.60, 11.07) ^***^ |  | 2.35 (1.52, 3.64) ^***^ |
| Exposure - Low G | 1.06 (0.59, 1.90) |  | 1.85 (0.82, 4.20) |  | 1.29 (0.69, 2.39) |
| Exposure - High G | 2.07 (1.30, 3.32) ^**^ |  | 9.68 (5.27, 17.78) ^***^ |  | 4.04 (2.47, 6.62) ^***^ |
| Multiplicative *P* value | **0.089** |  | 0.683 |  | 0.450 |
| RERI (95% CI) | 0.98 (-0.06, 2.06) |  | 2.52 (-1.60, 8.38) |  | 1.40 (-0.29, 3.50) |
| AP (95% CI)) | 0.47 (-0.05, 0.77) |  | 0.26 (-0.15, 0.50) |  | 0.35 (-0.10, 0.59) |
| **Exposure> weekly** |  |  |  |  |  |
| No exposure - Low G | 1.00 (reference) |  | 1.00 (reference) |  | 1.00 (reference) |
| No exposure - High G | 1.00 (0.67, 1.49) |  | 6.18 (3.64, 10.50) ^***^ |  | 2.50 (1.64, 3.83) ^***^ |
| Exposure - Low G | 0.92 (0.45, 1.87) |  | 1.65 (0.63, 4.28) |  | 1.52 (0.76, 3.03) |
| Exposure - High G | 2.48 (1.52, 4.06) ^**^ |  | 10.59 (5.77, 19.44) ^***^ |  | 4.63 (2.73, 7.83) ^***^ |
| Multiplicative *P* value | **0.021** |  | 0.942 |  | 0.646 |
| RERI (95% CI) | **1.57 (0.36, 3.00)** |  | 3.76 (-0.58, 10.71) |  | 1.6 (-0.55, 4.35) |
| AP (95% CI)) | **0.63 (0.14, 0.87)** |  | 0.35 (-0.07, 0.58) |  | 0.35 (-0.17, 0.6) |
| **Exposure>1 year** |  |  |  |  |  |
| No exposure - Low G | 1.00 (reference) |  | 1.00 (reference) |  | 1.00 (reference) |
| No exposure - High G | 1.04 (0.68, 1.58) |  | 6.35 (3.62, 11.14) ^***^ |  | 2.33 (1.51, 3.61) ^***^ |
| Exposure - Low G | 1.10 (0.61, 2.00) |  | 1.85 (0.79, 4.31) |  | 1.41 (0.76, 2.62) |
| Exposure - High G | 2.17 (1.35, 3.50) ^**^ |  | 10.22 (5.54, 18.84) ^***^ |  | 4.16 (2.51, 6.88) ^***^ |
| Multiplicative *P* value | **0.092** |  | 0.771 |  | 0.544 |
| RERI (95% CI) | **1.03 (0.03, 2.03)** |  | 3.02 (-1.01, 7.05) |  | 1.42 (-0.35, 3.18) |
| AP (95% CI)) | **0.47 (0.10, 0.85)** |  | 0.30 (-0.01, 0.6) |  | 0.34 (0.00, 0.68) |
| **Specific cleaning agents** |  |  |  |  |  |
| **Ammonia** |  |  |  |  |  |
| No exposure - Low G | 1.00 (reference) |  | 1.00 (reference) |  | 1.00 (reference) |
| No exposure - High G | 1.21 (0.84, 1.76) |  | 6.59 (4.01, 10.83) ^***^ |  | 2.67 (1.81, 3.94) ^***^ |
| Exposure - Low G | 1.54 (0.66, 3.58) |  | 3.20 (1.22, 8.38) ^*^ |  | 2.24 (0.99, 5.05) |
| Exposure - High G | 2.65 (1.43, 4.89) ^**^ |  | 12.28 (6.08, 24.81) ^***^ |  | 4.64 (2.43, 8.87) ^***^ |
| Multiplicative *P* value | 0.506 |  | 0.344 |  | 0.624 |
| RERI (95% CI) | 0.89 (-1.39, 3.10) |  | 3.5 (-3.63, 14.35) |  | 0.74 (-2.72, 4.72) |
| AP (95% CI)) | 0.34 (-0.73, 0.69) |  | 0.28 (-0.43, 0.58) |  | 0.16 (-0.86, 0.52) |
| **Bleach** |  |  |  |  |  |
| No exposure - Low G | 1.00 (reference) |  | 1.00 (reference) |  | 1.00 (reference) |
| No exposure - High G | 1.10 (0.73, 1.67) |  | 6.48 (3.70, 11.35) ^***^ |  | 2.33 (1.52, 3.59) ^***^ |
| Exposure - Low G | 1.22 (0.68, 2.18) |  | 2.04 (0.90, 4.64) |  | 1.29 (0.69, 2.43) |
| Exposure - High G | 2.13 (1.32, 3.45) |  | 10.22 (5.54, 18.88) ^***^ |  | 4.23 (2.57, 6.95) ^***^ |
| Multiplicative *P* value | 0.218 |  | 0.578 |  | 0.388 |
| RERI (95% CI) | 0.81 (-0.35, 1.97) |  | 2.70 (-1.67, 9.04) |  | 1.60 (-0.15, 3.86) |
| AP (95% CI)) | 0.38 (-0.21, 0.69) |  | 0.26 (-0.16, 0.50) |  | 0.38 (-0.06, 0.61) |

a There were 157 individuals with asthma onset in adulthood, and cases with childhood-onset asthma were excluded in the analysis.

Abbreviation: G: genetic risk measured by PRS. RERI, relative excess risk due to interaction (part of the total effect due to interaction); AP, proportion attributable to interaction (proportion of the combined effect due to interaction).

The PRS was dichotomized into high and low genetic risk groups based on the median value.

The logistic regression model was adjusted for age, sex, the first five genetic components, employment, the highest education level, household income, smoking status, and BMI.

A RERI or AP greater than zero indicated significant additive interaction (in bold).

^*^ *P* <0.05; ^**^*P*<0.01; ^***^*P*<0.001.

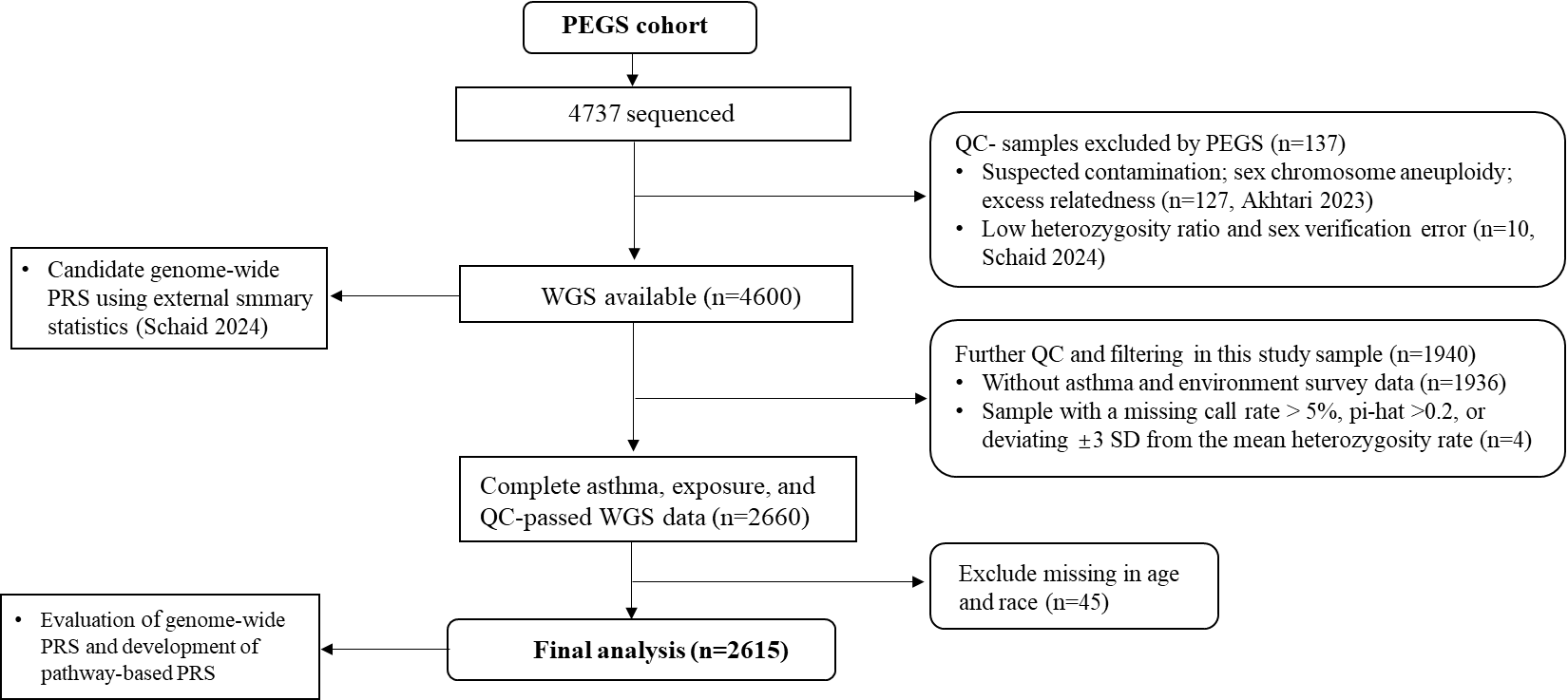

Figure S1 Sample processing, filtering and quality control for WGS data

Abbreviation: PEGS, Personalized Environment and Genes Study; WGS, whole-genome sequencing data; QC, quality control; PRS, polygenic risk score;

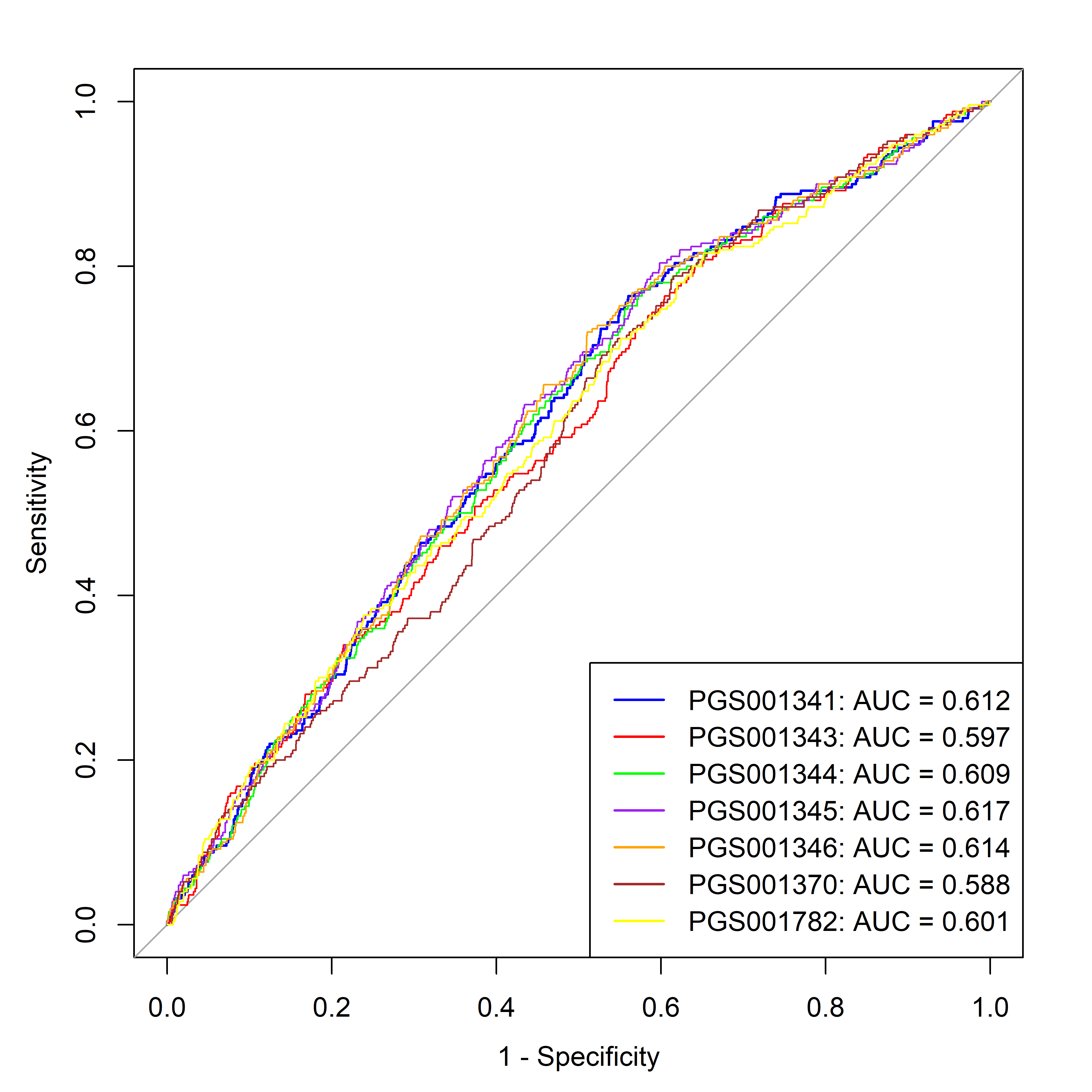

| PGSID | Total variants | Ambiguous variants | Non-auto variants | Used variants |
| --- | --- | --- | --- | --- |
| PGS001345 | 435 | 37 | 26 | 340/372 |

Figure S2 The ROC curves for PRS in the PEGS populations

The model included sex, age, first five genetic components, and each PRS.

Summary statistics of seven genome-wide PRS are from UK Biobank (Tanigawa Y, et al. doi: 10.1371/journal.pgen.1010105.) or Global Biobank Meta-analysis Initiative (Kristin Tsuo et al. doi: 10.1016/j.xgen.2022.100212; [Resources | Global Biobank Meta](https://www.globalbiobankmeta.org/resources)). Meta data were deposited in the PGS catalog. This study selected the best PRS (*PGS001345*) with the maximal AUC in the PEGS participants.

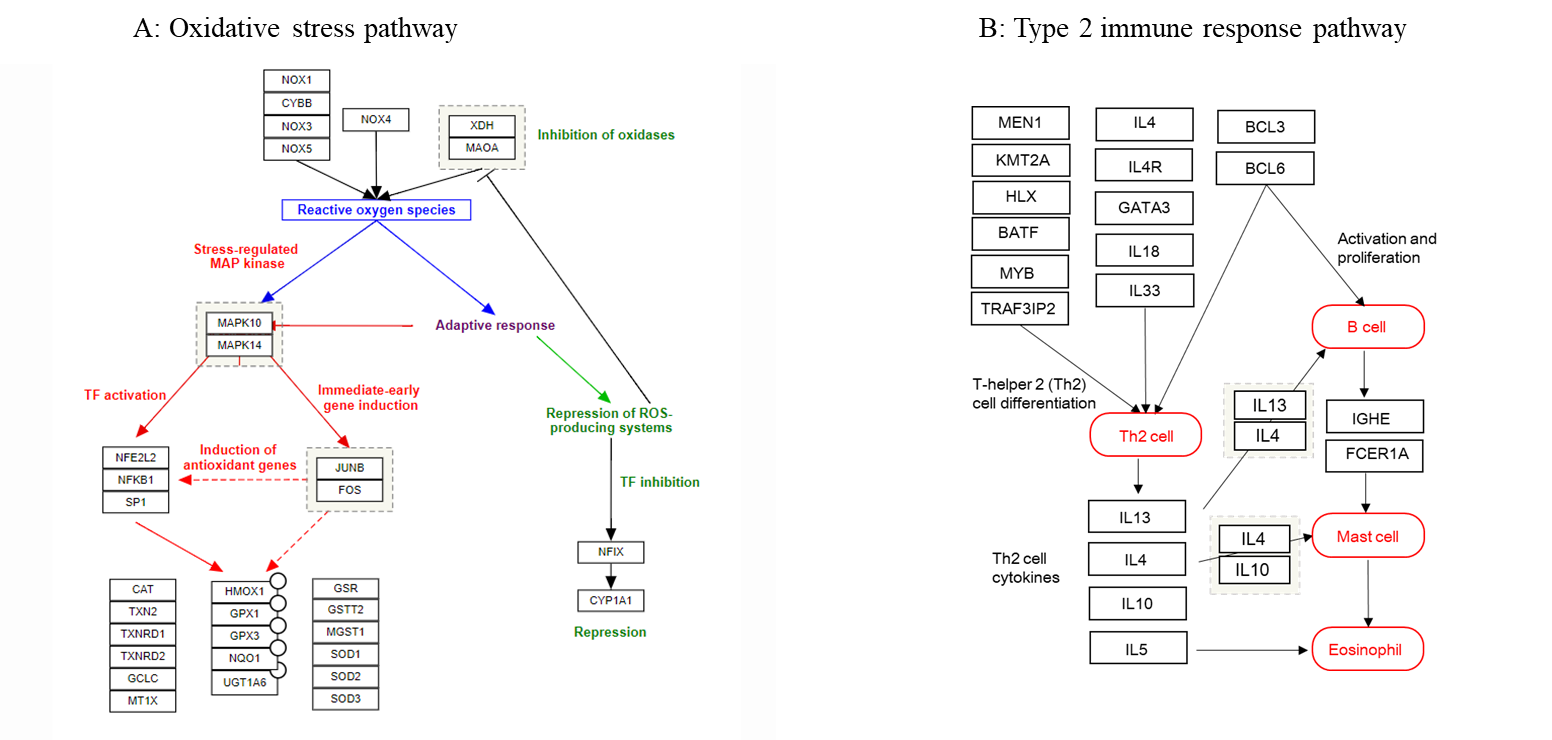
Figure S3 The gene sets involved in the hypothesized oxidative stress pathway and type 2 immune response

Gene set A: Curated and canonical WikiPathways ([WP408](https://www.wikipathways.org/pathways/WP408.html); Name: Oxidative stress response; Version: 20240227032141; Organism: Homo sapiens). No eligible biallelic SNP was found for GSTT2 within 1kb flanking area. Genes (CYBB, MAOA, and NOX1) located at sex chromosome were not included in the final analysis.

Gene set B: Adapted from Gene Ontology biological process; (GO:[0042092](https://www.gsea-msigdb.org/gsea/msigdb/human/geneset/GOBP_TYPE_2_IMMUNE_RESPONSE); Name: oxidative stress response; Filters: Organism-Homo sapiens; Access time: 2024-9-30) and KEGG pathway ([map05310](https://www.genome.jp/pathway/map05310), Name: Asthma; Version: 05310 8/17/20). No eligible biallelic SNP was found for IGHE and BCL6B within 1kb flanking area

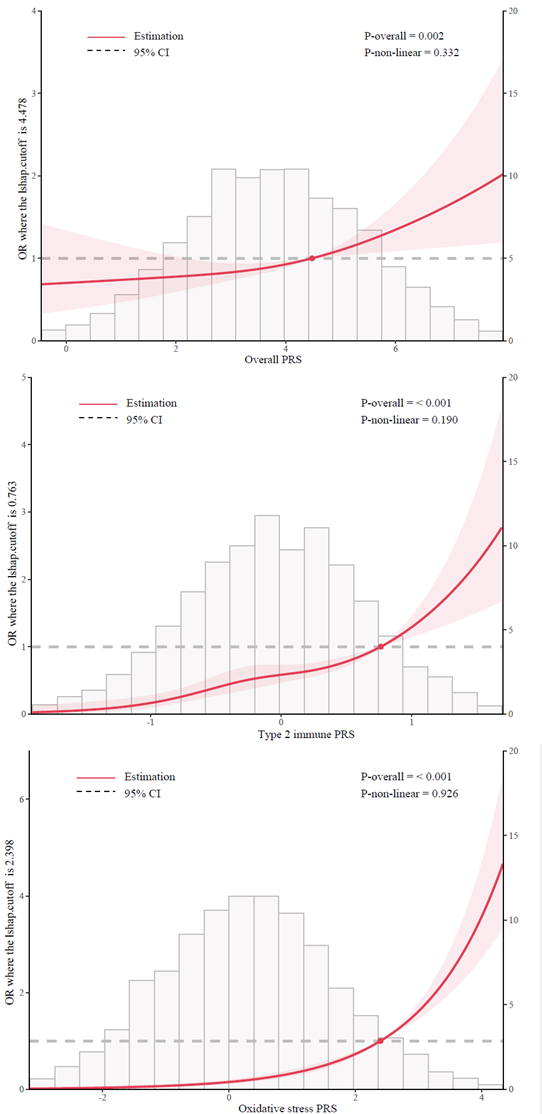
Figure S4 The relationship between overall and pathway-based PRS and adult asthma fitted by the restricted cubic spline.

Age, sex, and the first five genetic components were adjusted.

The number of knots was determined by the smallest BIC.

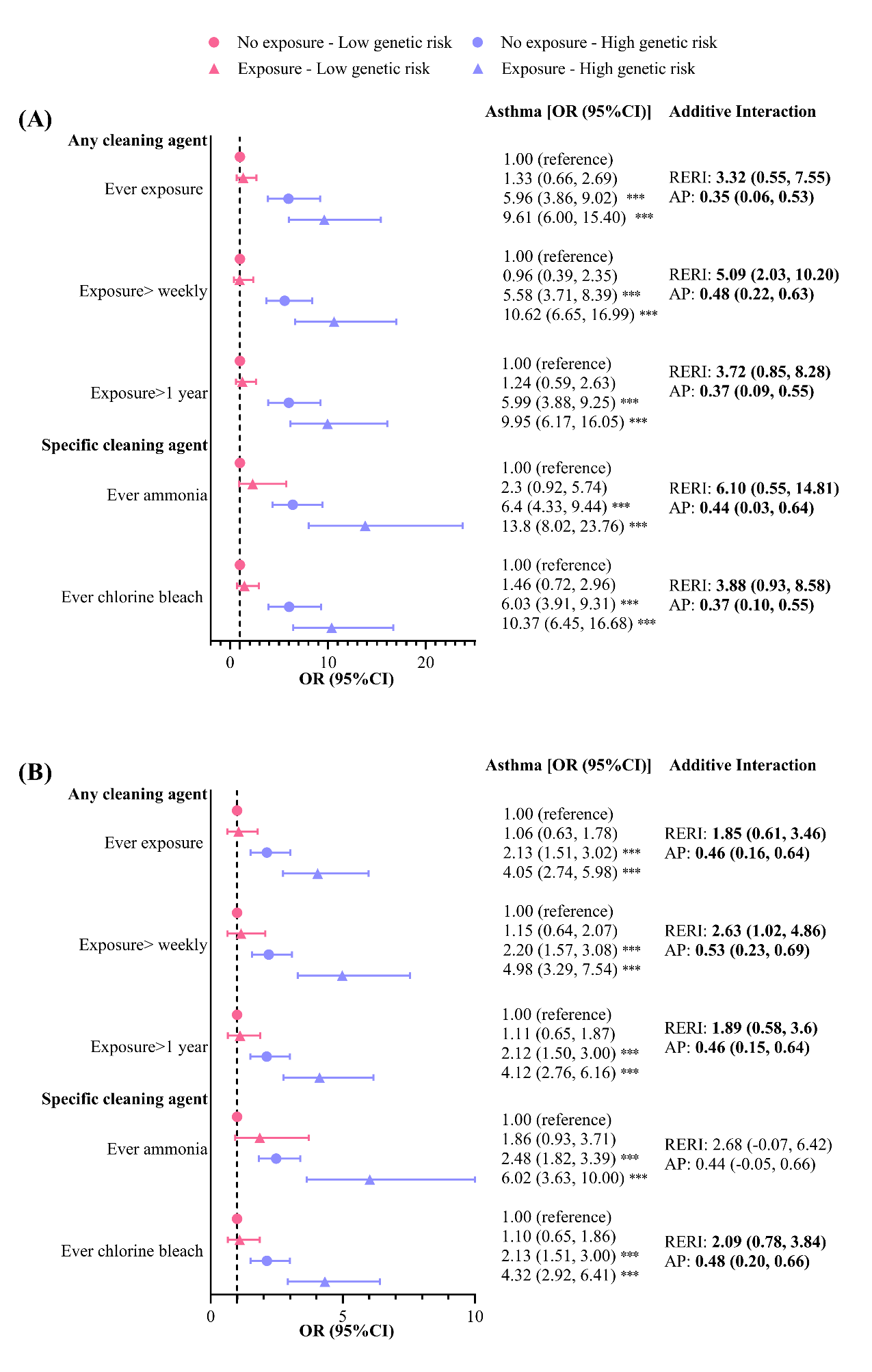
Figure S5 Sensitivity analysis of additive interaction of pathway-based PRS and occupational exposures on adult asthma with 2 kb extended gene regions (n=2615). (A) Oxidative stress pathway (B) Type 2 immune response

Abbreviation: RERI, relative excess risk due to interaction (part of the total effect due to interaction); AP, proportion attributable to interaction (proportion of the combined effect due to interaction).

The PRS was dichotomized into high and low genetic risk groups based on the median value.

The logistic regression model was adjusted for age, sex, the first five genetic components, employment, the highest education level, household income, smoking status, and BMI.

A RERI or AP greater than zero indicated significant additive interaction (in bold).

^*^ *P* <0.05; ^**^*P*<0.01; ^***^*P*<0.001.

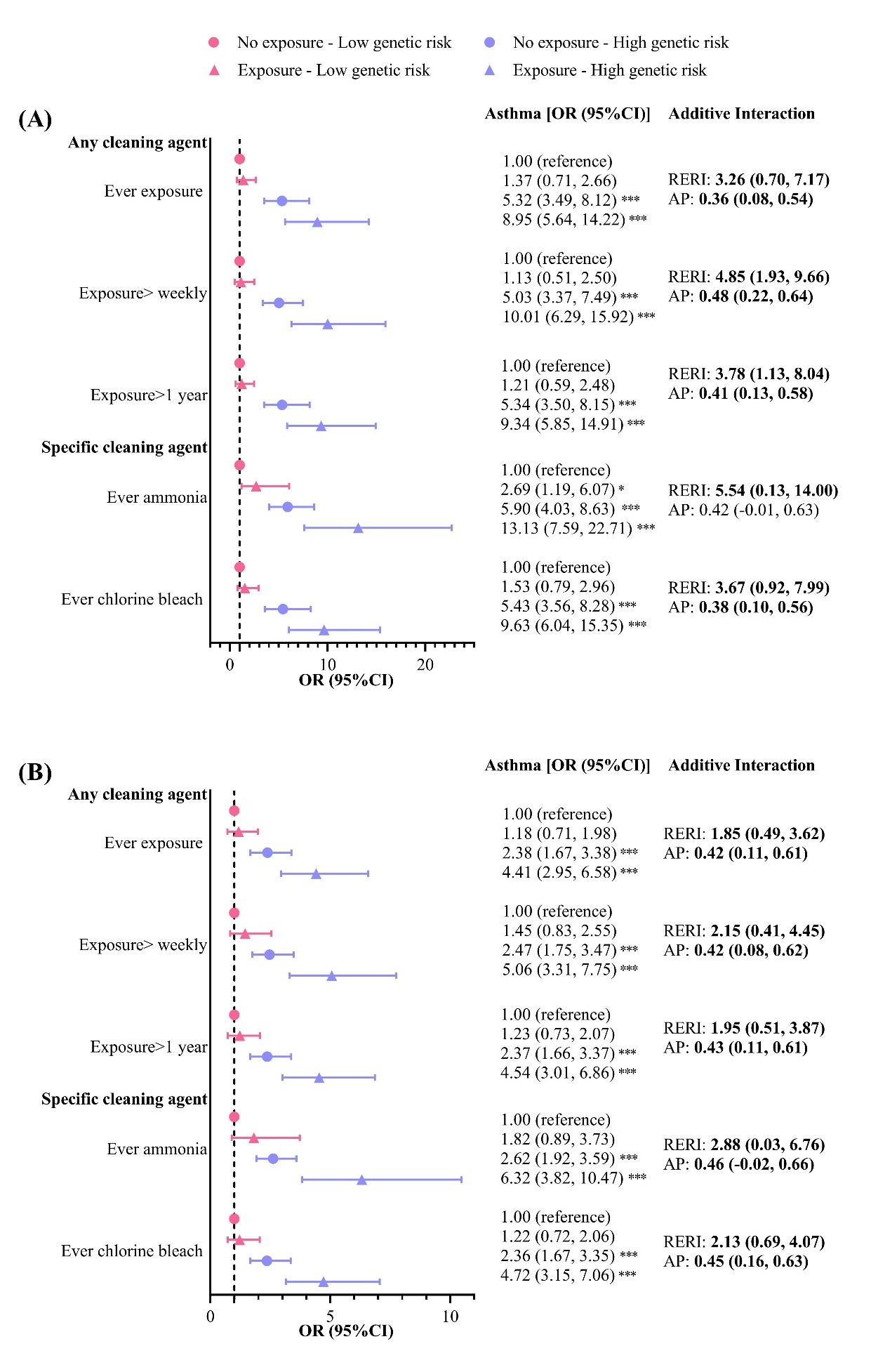
Figure S6 Sensitivity analysis of additive interaction of pathway-based PRS and occupational exposures on adult asthma with 5 kb extended gene regions (n=2615). (A) Oxidative stress pathway (B) Type 2 immune response

Abbreviation: RERI, relative excess risk due to interaction (part of the total effect due to interaction); AP, proportion attributable to interaction (proportion of the combined effect due to interaction).

The PRS was dichotomized into high and low genetic risk groups based on the median value.

The logistic regression model was adjusted for age, sex, the first five genetic components, employment, the highest education level, household income, smoking status, and BMI.

A RERI or AP greater than zero indicated significant additive interaction (in bold).

^*^ *P* <0.05; ^**^*P*<0.01; ^***^*P*<0.001.

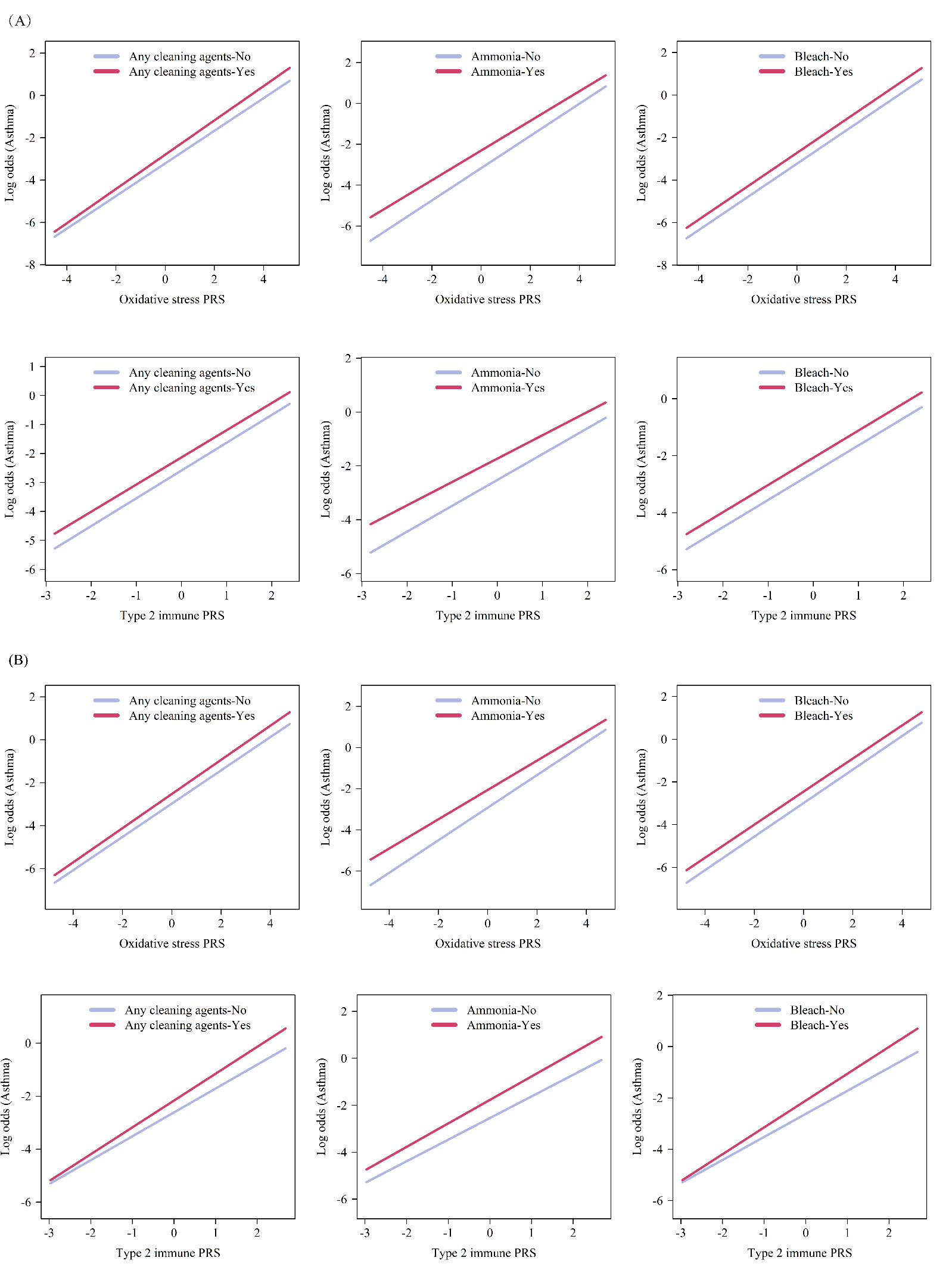

Figure S7 Sensitivity analysis of association between continuous pathway-based PRS and asthma modified by occupational cleaning agent exposure with different extended gene regions (n=2615)

(A) Gene annotation within 2 kb flanking area (B) Gene annotation with 5 kb flanking area

Abbreviations: PRS, polygenic risk score

The multiplicative interaction model was adjusted for age, sex, the first five genetic components, employment, the highest education level, household income, smoking, and BMI
